# Standardizing Nasal Fluid Processing for Respiratory Biomarker Analysis: A Comparative Study of Homogenization and Storage Methods

**DOI:** 10.1101/2025.06.11.25329425

**Authors:** Tanya Lupancu, Carley Xia, Eldin Rostom, David M. Yen, Brian Wang, Adam M. Damry

**Affiliations:** Diag-Nose Medical, Notting Hill, 3168, Victoria, Australia; Monash University, Clayton, 3800, Victoria, Australia; Specialty Physician Associates, Bethlehem, 18017, Pennsylvania, United States; University of Ottawa, Department of Chemistry and Biomolecular Sciences, Ottawa, K1N 6N5, Ontario, Canada

## Abstract

**Background:** Nasal fluid is a valuable medium for biomarker analysis in respiratory diseases due to its accessibility and proximity to relevant pathophysiological processes. However, its heterogeneous nature presents challenges for consistent biomarker detection and quantification. This study aimed to improve protein biomarker recovery and reliability by evaluating different homogenization and storage methods for nasal fluid samples.

**Methods:** Mechanical disruption techniques using syringes and commercial BioMasher units were compared for their effectiveness in analyte recovery, and the impact of pre- and post-processing of cryogenically stored nasal samples was assessed. Inflammatory protein biomarkers relevant to respiratory diseases, including interleukin (IL)-13, IL-8/ CXCL8, myeloperoxidase (MPO), CCL11/ eotaxin-1, CCL26/ eotaxin-3, elastase, myxovirus resistance protein 1 (MxA), CCL17/ TARC, CXCL10/ IP-10), and serum amyloid A (SAA), were successfully detected in the processed nasal fluid by ELISA and Luminex assays.

**Results:** Significant differences in analyte recovery were observed between homogenization methods, with greater titers of IL-5 and eotaxin-1 measured in samples processed by the syringing method. However, homogenization could be performed either before or after long-term cryogenic storage without significantly affecting protein concentrations. Spike and recovery experiments were also conducted and differences in SAA protein recovery were detected, suggesting that the matrix effects in nasal fluid can affect cytokine recovery, but variations in recovery were not significant for other biomarkers tested.

**Conclusions:** These findings demonstrate the potential of nasal fluid as a readily accessible medium with substantial potential to aid diagnosis and monitoring of respiratory diseases. However, given the significant impact homogenization protocols had on analyte recovery, the growing use of nasal fluid as a biospecimen in the respiratory field highlights the critical need for standardized protocol to ensure accuracy and reproducibility in future analyses.

## INTRODUCTION

Nasal fluid, also referred to as nasal mucus, is a gel-like substance that coats the respiratory epithelium. It has both functional and protective roles in the respiratory system, maintaining nasal passage moisture to facilitate airflow, while trapping foreign particles and pathogens to protect the lungs. Mucins are a major component of mucus and the hydration of these large bottlebrush-shaped glycoproteins determine mucus viscosity (1, 2). In addition to mucins, nasal fluid contains immunomodulatory proteins such as antibodies and cytokines that interact with pathogens, allergens, and environmental factors to facilitate immune recognition and initiate appropriate inflammatory responses (3, 4). Amongst these, cytokines regulate inflammation and activate immune cells, contributing to the protection of the upper respiratory tract from infections and irritants (5, 6). IgE antibodies similarly mediate the immune system’s response to environmental allergens and play a central role in allergic reactions, with elevated levels detected in the nasal lavage fluid of patients with allergic rhinitis (7, 8). Given the role of these immunomodulators in pathological response, variations in protein concentrations can serve as biomarkers for these conditions, making nasal fluid a valuable medium for disease detection, diagnosis, and monitoring (6, 9, 10).

Asthma is the most common chronic respiratory disease and it is characterised by elevated levels of type 2 cytokines such as IL-4, IL-5 and IL-13, which drive key pathological features of the disease such as mucus overproduction, hyperresponsiveness, and airway eosinophilia and remodelling (5, 6, 11). As contributors to asthma pathogenesis, these cytokines are therapeutic targets and treatments are designed to either reduce their expression or inhibit their downstream activity. Oral corticosteroids (OCS) are commonly prescribed for asthma management as they can quickly reduce inflammation and associated symptoms; however, their long-term use is associated with adverse effects due to the widespread distribution of the glucocorticoid receptor and subsequent non-specific immunosuppression (12). Biologics offer a more targeted approach to immunosuppression as they inhibit the activity or signalling pathways of select immunological proteins. Dupilumab (Dupixent, Regeneron) is a human monoclonal antibody that binds to the alpha subunit of the IL-4 receptor, thereby blocking the downstream signalling pathways activated by IL-4 and IL-13 (13–15). While the drug is prescribed to asthmatics with evidence of type 2 inflammation, this broad classification does not successfully capture the heterogenous nature of asthma (16).

Notably, while type 2 inflammation in patients can result from elevated levels of various type 2 cytokines, inflammation is instead broadly categorized by the quantification of biomarkers such as elevated blood eosinophils, fractionated exhaled nitric oxide (FeNO), and IgE (17). As a result, the specific cytokines responsible for the disease are not precisely identified. Given the availability of other biologics targeting other proinflammatory factors, including the IL-5 antagonist mepolizumab (Nucala, GSK), the TSLP antagonist Tezepelumab (Tezspire, Amgen/AstraZeneca), and the IgE antagonist omalizumab (Xolair, Novartis), the exclusion of specific cytokine profiles from treatment criteria thus limits the potential for precision medicine in asthma management (18). Moreover, type 2 inflammation may be the predominant phenotype in asthma but a subset of asthmatics have non- or low-type 2 inflammation, which is even less responsive to standard biologic therapies (19).

Neutrophil-associated proteins such as IL-8, myeloperoxidase and elastase can be detected at elevated levels in airway of these individuals (20–23). Only a select few of these targets are incorporated into disease management strategies, leading to a lack of precision that likely contributes to the challenge of treating non-responders (24).

As a biospecimen that reflects the nasal environment, nasal fluid provides an opportunity to overcome this diagnostic barrier. Profiling of nasal fluid samples has traditionally been used to identify pathogens responsible for respiratory infections, such as the SARS-CoV-2 virus for COVID-19 diagnosis. Proteomic profiling of this fluid could also reflect underlying changes in the respiratory system to enable clinical decision-making and tailored treatment approaches (25). Such analyses, however, are complicated by the heterogeneous nature of the nasal fluid medium. Nasal fluid samples can vary widely in viscosity and composition, and thus require processing steps to obtain a homogeneous and stable sample (3). However, current protocols for the collection, processing, and storage of these heterogeneous samples are inconsistent, leading to high study-to-study variability (26). Repeated freeze/thaw cycles, due to improper sample handling and aliquoting, can additionally increase both protein aggregation and degradation, introducing further error in downstream quantitation (27–29). The matrix effects of nasal fluid, caused by mucins, endogenous proteases, and other biological components present, can also affect the accuracy and sensitivity of immunoassays and mass spectrometry-based analyses (30, 31). Altogether, these factors highlight the need for standardized protocols to ensure the reproducibility and reliability of nasal fluid biomarker quantification.

In this study, we focused on understanding sources of variability in nasal fluid processing, focusing on nasal fluid collected via nasal blowing, which requires minimal specialized equipment. For sample processing and homogenization, we compared the mechanical shearing capability of syringe aspiration with the commercially available BioMasher I homogenization kit. The performance of these methods was evaluated by quantification of the recovery of key immune biomarkers from these samples. We also verified the impact of ordering of processing steps and of the nasal matrix on cytokine recovery to better establish a reliable protocol for future clinical and research purposes.

## MATERIALS AND METHODS

### Sample Collection

The nasal samples used in this research were from Lee BioSolutions Inc (Cat # 991-13-S), a major biomedical database. Due to low sample volume, different lot numbers were used between experiments. In the sample homogenisation study, Sample 1 (Donor T6445; Lot # 20-10-548), Sample 2 (Donor T6672; Lot # 21-01-716), Sample 3 (Donor T6303; Lot # 21-01-586) and Sample 4 (Donor T5631; Lot # 21-02-654) were all from Lee BioSolutions. In the spike and recovery study, Sample 1 (Donor T6521; Lot # 20-07-621), Sample 2 (Donor T6675; Lot # 20-09-598), Sample 3 (Donor T6672; Lot # 20-10-634) and Sample 4 (Donor T6292; Lot # 21-01-532) were from Lee BioSolutions.

In the stability study, Sample 1 (Donor T6343; Lot # 21-02-554), Sample 2 (Donor T6557; Lot # 20-12-753), Sample 3 (Donor T6406; Lot # 21-01-510) and Sample 4 (Donor T6655; Lot # 20-09-645)

### Sample Homogenization

Nasal samples were homogenized using two different methods: mechanical disruption with a syringe (“syringing”) and with a BioMasher I. For syringing, nasal samples were aspirated 10 times through a 23-gauge syringe, followed by centrifugation at 20,000g for 30 minutes at 4°C. The second method used the BioMasher I with an O-ring (Cat # NIP-30-1.5-O, Nippi Inc), as per manufacturer’s instructions. Briefly, 125μL of unprocessed nasal sample was added to a filter tube and homogenized with a pestle equipped with an O-ring, followed by centrifugation at 15,000g for 30 seconds. The supernatant from both homogenisation methods was collected and used for downstream analysis.

### Spike and Recovery

Nasal fluid was spiked with a high and low concentration of protein to assess the impact of the nasal fluid matrix on protein recovery (Supplementary Table 1). Proteins included recombinant human CCL17/TARC protein (Cat # 364DN, R&D Systems), recombinant human elastase protein (Cat # 230-00693-10, RayBiotech), recombinant human IL-8/CXCL8 protein (Cat # 230-00010-10, RayBiotech), recombinant human CXCL10/ IP-10 protein (Cat # 266-IP, R&D Systems), recombinant human Serum Amyloid A1 protein (NBP2-61366, Novus Biologicals) and MxA Protein Human E.coli (Cat # RD172349100, BioVendor R&D).

An eight-point standard curve for each protein determined the spiking concentrations, with the high concentration set at 80% of the first standard curve value, and the low concentration set at 80% of the value between points 4 and 5 from the top of the standard curve.

### Protein Quantification

Nasal samples were diluted 100-fold in assay buffer before total protein levels were measured using the Qubit Protein Assay kit (Cat # 33212, Thermofisher) on a Qubit 4 Fluorometer (Thermofisher). Urea levels in the nasal fluid samples were measured using the Urea Colormetric Assay Kit (Cat # K375-100, BioVision, Inc), as per manufacturer’s instructions.

Free haemoglobin concentrations were measured using the Harboe method, with absorbances recorded at 380nm, 415nm and 450nm. The values were applied to the following formula for quantification: Free hemoglobin (µg/mL) = 836 × (2 × A415nm − A380nm − A450nm) × 1000. A standard was generated using Hemoglobin Human (Cat # H7379, Sigma-Aldrich).

### ELISA

Human MxA ELISA kit (Cat # RD194349200R, BioVendor), human IL-8 ELISA kit (Cat # ELH-IL8-2, RayBiotech), human TARC ELISA kit (Cat # ELH_TARC-1, RayBiotech) and human elastase ELISA kit (Cat # HK319, Hycult Biotech) were used to measure proteins levels of MxA, IL-8, CCL17 and Elastase, respectively. Protein levels were measured by ELISA on a Tecan Infinite F50 (Tecan Group).

### Luminex

CCL17, IP-10 and SAA protein levels were measured using a custom 3-plex Luminex assay (Cat # PPX-03-MX47XAR). Eotaxin-1, IL-13, IL-5, IL-8 and MPO were measured using a custom 5-plex Luminex assay (Cat # PPX-05-MXCE47D) and eotaxin-3, IL-13, IL-5, IL-8 and MPO (Cat # PPX-05-MXAACUT) were measured on another custom 5-plex Luminex assays. These assays were sourced from Thermofisher and run on a MAGPIX System running xPONENT (ThermoFisher), as per manufacturer’s instructions. Protein results were analysed on Belysa Analysis Software (Millipore Sigma).

### Statistical Analysis

Statistical analyses were performed using unpaired t-test for comparing mean of two independent groups, paired t-test for comparing mean of two dependent groups, or two-way ANOVA with Sidak post-test for more than two different groups, as indicated. A p value < 0.05 indicates significance. Data were graphed using GraphPad Prism version 9.5.1.

## RESULTS

### Comparison of nasal samples homogenization methods

The viscosity of bulk nasal samples between patients can be highly variable, with the method of homogenization used potentially influencing the result of downstream analyte analyses. Our aim was to compare different homogenization protocols and evaluate their effect on quantifiable analyte levels in the resulting processed nasal samples. Tissue samples can be homogenized via mechanical shearing through a narrow aperture, such as, for example, through repeat needle aspiration.

Commercial homogenizations kits are also available for easy protein and nucleic acid extraction from tissue samples (32). We compared the efficacy of these two homogenization techniques on our nasal samples. Total protein, urea and haemoglobin levels were measured, with no significant differences observed between the different methods (Figure 1A-C). These basic controls are a metric of sample integrity and recovery, and neither method retained biological material to a significantly greater extent. In addition, while no observable blood contamination could be observed visually (as a discoloration of the sample) one sample demonstrated a substantially higher level of detected hemoglobin relative to the other three. This elevated hemoglobin concentration is potentially indicative of the presence of dilute blood contamination, although this was observed across both processing methods, and therefore did not impact comparative analyses.

**Figure 1:**
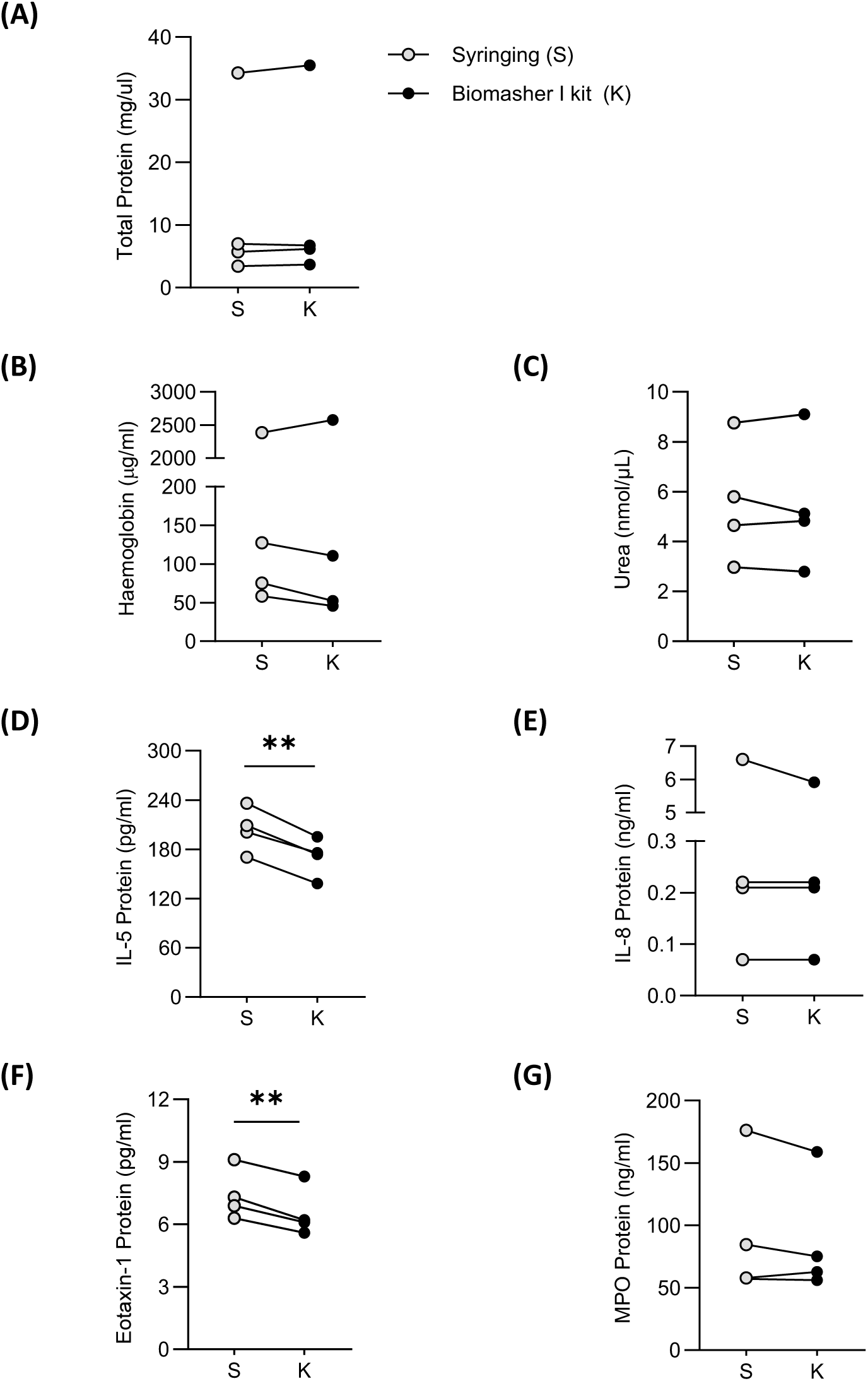
Method of Nasal Sample Homogenization Affects Cytokine Concentration. Bulk nasal samples were homogenized via syringing (S) or the BioMasher I kit (K). (A) Total protein, hemoglobin, (C) urea, (D) IL-5, (E), IL-8, (F) eotaxin-1/ CCL11 and (G) and MPO protein concentrations were measured and compared. The data is graphed as dot plots, n=4, and p values were obtained using paired T-test, **p<0.01.

We next investigated whether the recovery of specific protein biomarkers differed between the two methods. The homogenized sample concentrations of proteins of varying sizes were measured, including IL-5 (13.3 kDa, dimeric), IL-8 (also known as CXCL8, 8.9 kDa), eotaxin-1 (also known as CCL11, 8.4 kDa) and myeloperoxidase (MPO, 84 kDa, dimeric). IL-5 and eotaxin-1 protein recovery was significantly higher in samples processed via the syringing method; IL-5 recovery increased 20%, with average protein concentrations increasing from 170pg/ml to 204pg/ml. and Eotaxin-1 recovery increased 13%, with average protein concentrations increasing from 6.55pg/ml to 7.4pg/ml (Figure 1D, E). In contrast, IL-8 and MPO levels were not substantially different between both homogenisation techniques (Figure 1F, G). Syringing was reported to be less labour-intensive due to the reduced number of procedural steps, and it was also identified as a more cost-effective method. Given this method’s equivalent to greater performance for analyte recovery relative to the BioMasher I across the set of proteins tested, the syringing method was used in subsequent experiments.

### Effects of sample storage on biomarker measurements

We next evaluated whether the freezing of nasal samples, whether prior to or following sample processing, affected sample integrity and biomarker recovery. Four samples of increasing viscosity and decreasing clarity were chosen to verify freezing effects over a representative subset of samples. These samples were aliquoted and either frozen neat or post-homogenisation via syringing for up to 56 days. The total protein concentration showed minimal variation across either the duration of cryogenic storage or the order of homogenisation and storage, highlighting that the bulk matrix properties of the nasal mucus samples are not subject to substantial variation under these conditions (Figure 2A). The detectability of selected immune biomarkers (IL-5, IL-8, eotaxin-3, and MPO) in ELISAs was also assessed (Figure 2B-E). No significant differences in biomarker concentration were observed relative to the order of processing and freezing. Variability over the storage duration was greater than for total protein concentration measurement, but no distinct trends were observed for any biomarker. This result indicates that sample viability for testing is maintained over prolonged storage times under cryogenic conditions whether or not samples have been homogenised.

**Figure 2:**
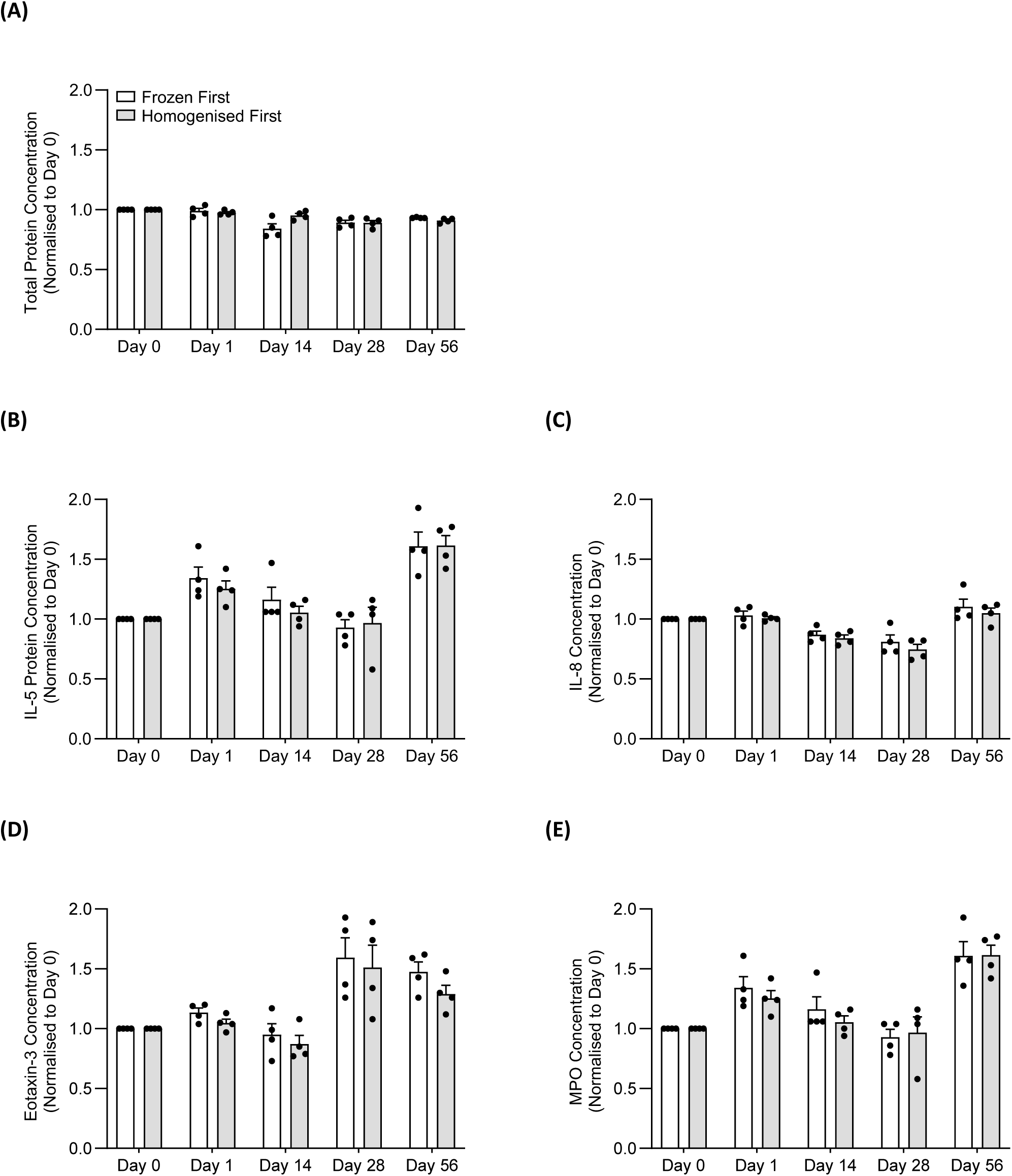
Cytokine Concentration in Nasal Samples is Unaffected by Processing Order. Bulk nasal samples were frozen for up to 56 days, with homogenization performed either before or after freezing. (A-E) Protein concentrations were measured and normalized to day 0. The data is graphed as scatterplots with bars indicating mean ± SEM, n=4. 2-way ANOVA was applied, though no statistically significant difference was observed between the processes.

### Matrix effects in homogenized nasal fluid

Spike and recovery experiments were next conducted to assess whether the complex matrix of nasal fluid affected biomarker recovery. Four samples of varying viscosities were spiked with low or high protein concentrations (PC) of a range of selected biomarkers with differing molecular properties, including CCL17 (also known as TARC), elastase, IL-8, IP-10 (also known as CXCL10), serum amyloid A (SAA) and human myxovirus resistance protein-1 (MxA). Prior to spike and recovery measurements, baseline concentrations of these biomarkers in each sample were measured to establish appropriate spike PCs (Supplementary Table 1) and dilution factors (Supplementary Table 2). These baseline protein measurements varied substantially between samples, and no correlations to viscosity or cross-correlations to other biomarker concentrations were observed (Supplementary Table 3). CCL17 protein concentrations varied over 15-fold between the four samples, elastase-1 varied 60-fold, IL-8 and IP-10 varied over 21- and 25-fold, respectively, SAA varied over 1000-fold and MxA varied 45-fold.

The protein recovery of these spiked samples was measured and calculated as a percentage of theoretical concentration (Figure 3 and Supplementary Table 3). CCL17 recovery averaged 72.1% in low-PC samples and 82.6% in the high-PC samples, with significant variation observed. Elastase, IL-8, and IP-10 protein recoveries were high and consistent across samples, averaging greater than 90% recovery in both low- and high-PC samples with no statistical differences between these samples. MxA recovery was similarly not statistically different between low- and high-PC samples, but ranged to lower average recoveries, especially in the high-PC sample, where an average recovery of 84% was observed. SAA protein recovery was statistically different between the spiked samples, with 93.4% and 79.4% of SAA recovered in the low- and high-PC samples, respectively. Descriptive statistics of these recovery percentages, including coefficients of variation, are outlined in Supplementary Table 3. These results suggest that matrix effects in nasal fluid may affect cytokine recovery, though overall recovery remained high across different proteins and concentrations.

**Figure 3:**
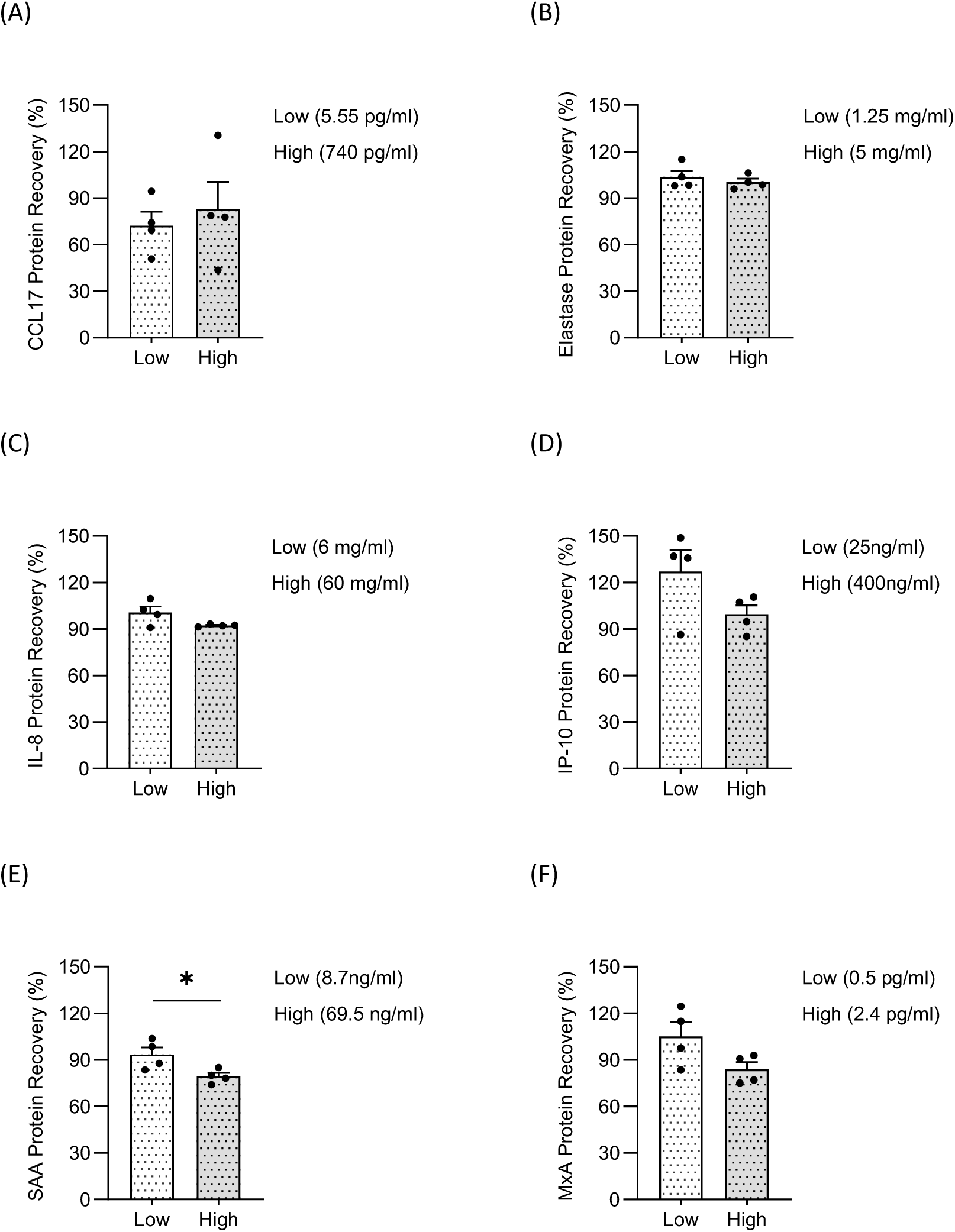
Nasal fluid matrix does not affect protein recovery. Processed samples were spiked with low or high protein concentrations. (A-F) Protein recovery was calculated as measured protein concentration ÷ spiked protein concentration x100. The data is graphed as scatterplots with bars indicating mean ± SEM, n=4. p values were obtained using paired T-test, *p<0.05.

## DISCUSSION

While nasal fluid is a rich and dynamic source of biomarkers that report on respiratory and systemic health, it is also known for its heterogeneous physical composition, presenting significant challenges for consistent biomarker detection and quantification. Although proper handling of nasal fluid samples is critical for optimal analyte recovery and ensuring reproducibility across samplings, current research has predominantly focused on optimizing sampling techniques and standardized methods for handling of these samples have yet to be established (26, 33). By evaluating different sample homogenization and processing methods, this research aims to refine basic protocols that can improve the reproducibility and reliability of nasal fluid analyses.

Although homogenization is widely used across biological sample types, including methods such as enzymatic digestion, chemical treatment, and sonication, the optimal approach depends on the intended analysis. Here, we sought to establish a standard for nasal fluid sample processing that required minimal specialized equipment. In this work, we compared two simple and easily accessible homogenization methods through mechanical disruption using either a standard syringe or the BioMasher I. Our results demonstrated that both methods led to similar sample integrities, as measured by total protein content, but that syringing was superior to the BioMasher I for cytokine recovery. Although both methods appeared to reach homogeneity, this highlights that disruption of the sample is likely to be different across methods. It is also possible that the plastics used in the BioMasher I may additionally adsorb a greater range of low-concentration analytes that are more conserved in the sample by syringing, as it is well established that many cytokines will readily and differentially adsorb to various plastics (34). More broadly, these findings reinforce this principle that cytokine recovery varied by homogenization method, whereas total protein concentration remained unaffected, underscoring the need for a standardized protocol. Given its lower labour demands and cost-effectiveness, along with an equal-to-better recovery of tested biomarkers, we recommend establishing a standard syringing protocol for routine laboratory use.

In addition to homogenization, there is also a lack of standardization regarding how nasal fluid is stored. While cryogenic storage is common for the transportation and long-term storage of most if not all biospecimens to mitigate degradation, the impact of freezing these samples, whether before or after homogenization, is equally poorly understood (35). Nonetheless, as it is known that levels of cytokines and other biomarkers in other biospecimens such as plasma can be adversely affected by freezing, it is important to establish standard methodologies to minimize variability in downstream sample measurements (29, 36). Here, we observed no significant decreases in total protein content or in the concentrations of any tested biomarker over roughly two months of cryogenic storage. This result differs from previous studies that have shown significant decreases in IL-8 and MPO concentrations in nasal lavage samples after long-term cryogenic storage (37). In addition, it is common practice to store unprocessed biospecimens as the high biomolecular density of unprocessed samples generally provides a cryoprotecting effect to biomarkers within the sample (36). However, our results indicated little difference in biomarker concentrations whether samples were frozen unprocessed or homogenized. These two results together suggest that the mucin-rich environment of nasal fluid likely mimics this cryoprotectant effect regardless of the order of freezing and processing. Even so, it should be noted that this conclusion is limited to the specific biomarkers and timeframe examined, as unmeasured analytes may be more susceptible to degradation.

The complex nasal fluid matrix may also present substantial disadvantages for analyte quantification through non-specific matrix effects. These effects, which are commonly observed when handling minimally diluted biological fluids, can lead to substantial variability in recovery rates (38). Our spike-and-recovery experiments in homogenized nasal fluid revealed low-level matrix effects overall, with most tested proteins exhibiting recoveries of >80%. Nonetheless, recoveries did vary by biomarker identity and concentration, hinting that matrix effects are affecting measurement linearity in these samples. Similar effects are observed in plasma and serum, where cytokine levels differ due to differential platelet and leukocyte activation during processing (39, 40). Although our samples showed no visible blood contamination, the nasal environment is highly dynamic, and moreover, nasal fluid contains variable cellular populations depending on collection method (e.g., passive wicking vs. scraping) (26, 41). Therefore, it is important to understand signal interference caused by the nasal fluid matrix, and to validate at what dilutions these become insignificant, restoring linearity during quantification. As matrix effects are typically caused by interfering substances in the matrix, these could also be targeted specifically for removal or attenuation. Albumin, a major plasma protein also present in nasal fluid, binds small molecules and can alter their measured concentrations (32, 42, 43). Additionally, mucins, highly glycosylated proteins abundant in nasal secretions, can interact with cytokines and other proteins, potentially affecting recovery rates (44). Despite these early-stage hurdles complicating its use as an analytical biospecimen, nasal fluid is nevertheless an easily accessible and non-invasive biofluid that holds great promise for respiratory biomarker research. Its composition reflects local and systemic inflammatory processes, making it a valuable medium for disease monitoring. However, variability in sample handling and processing can significantly impact analyte detection. Limitations of this study include a small sample size and a restricted cytokine panel, nonetheless, our findings highlight the importance of standardizing nasal fluid processing to improve analytical reproducibility. The proposed protocol (Figure 4) provides a framework for bulk nasal fluid homogenization, optimizing cytokine analysis while minimizing the need for specialized equipment. In parallel, standardizing collection methods will further enhance consistency across studies. By adopting standardized protocols, nasal fluid research can advance respiratory biomarker discovery, ultimately supporting more precise and patient-centered diagnostics.

**Figure 4:**
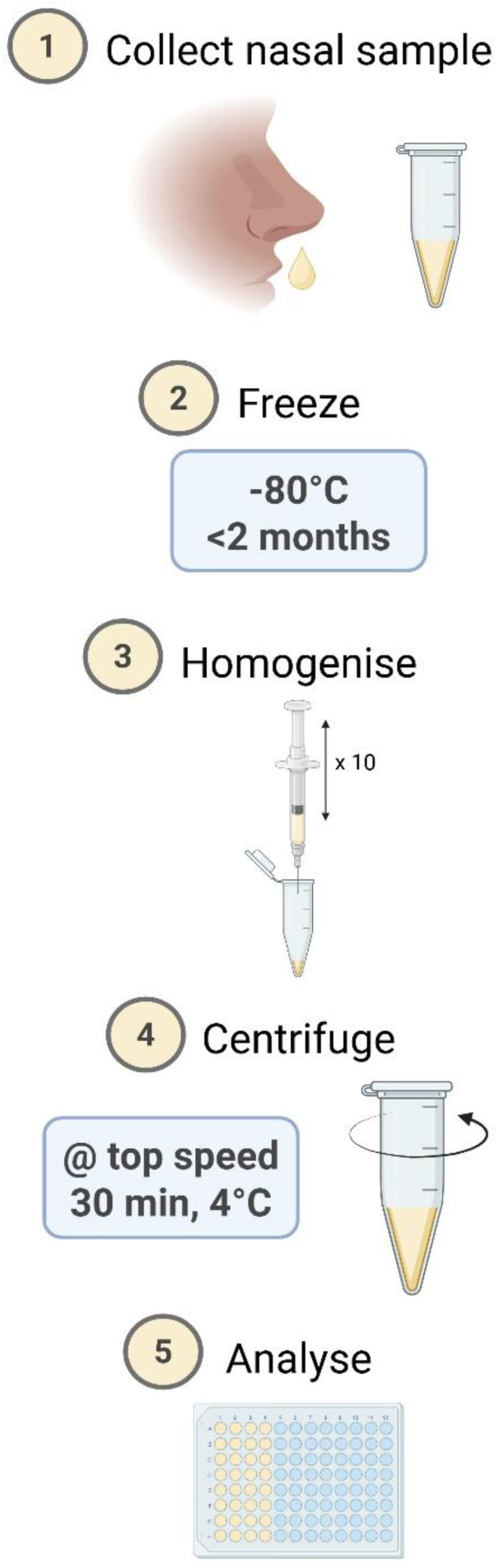
Suggested protocol for bulk nasal sample preparation for downstream cytokine analysis. Bulk nasal fluid is a highly heterogeneous medium rich in analytes of interest. To maintain sample integrity, it is recommended to store samples at -80°C for up to 2 months post-collection. For analysis, samples should be homogenized by simple syringing, then centrifuged at top speed (16,000–20,000g) for 30 minutes at 4°C. The supernatant is then suitable for cytokine analysis via single- or multiplex assays. Created in BioRender. Lupancu, T. (2025), https://BioRender.com/w4u4p8k

## Data Availability

All data generated or analysed during this study are included in this published article and its supplementary information files.

## LIST OF ABBREVIATIONS

ANOVA: Analysis of Variance
Cat #: Catalogue Number
CCL11: C-C Motif Chemokine Ligand 11
CCL17: C-C Motif Chemokine Ligand 17
CCL26: C-C Motif Chemokine Ligand 26
COVID-19: Coronavirus Disease 2019
CXCL8: C-X-C Motif Chemokine Ligand 8
CXCL10: C-X-C Motif Chemokine Ligand 10
ELISA: Enzyme-Linked Immunosorbent Assay
GSK: GlaxoSmithKline
IL-13: Interleukin-13
IL-8: Interleukin-8
IP-10: Interferon Gamma-Induced Protein 10
MPO: Myeloperoxidase
MxA: Myxovirus Resistance Protein A
PC: Protein Concentration
SAA: Serum Amyloid A
SARS-CoV-2: Severe Acute Respiratory Syndrome Coronavirus 2
SPA: Specialty Physician Associates
TARC: Thymus and Activation-Regulated Chemokine

## DECLARATIONS

### Ethics approval and consent to participate

Not applicable

### Consent for publication

Not applicable

### Authors’ contributions

TL conducted the formal analysis and contributed to conceptualisation, review and editing of the manuscript. CX wrote the original draft. AD contributed to the conceptualization and design of the study and participated in manuscript review and editing. ER, DY, and BW contributed to the conceptualization and study design. All authors read and approved the final manuscript.

## Acknowledgements

We thank L. Kaur and A. Yusim of Crux Biolabs for performing the experiments.

