## Supplementary Tables 1-9 for "Standardizing Nasal Fluid Processing for Respiratory Biomarker Analysis: A Comparative Study of Homogenization and Storage Methods"

**Supplementary Table 1: Low and high protein concentration of cytokines used for spiking experiment.**

| Cytokine | Low | High |
|---|---|---|
| **CCL17 (pg/ml)** | 5.55 | 740 |
| **Elastase-1 (mg/ml)** | 1.25 | 5 |
| **IL-8 (mg/ml)** | 6 | 60 |
| **IP-10 (ng/ml)** | 25 | 400 |
| **SAA (ng/ml)** | 8.69 | 69.5 |
| **MxA (pg/ml)** | 0.48 | 2.4 |

**Supplementary Table 2: Cytokine concentrations and dilution factor of neat nasal samples before being spiked with low or high concentrations of these cytokines.**

| Cytokine | Sample 1 | Sample 2 | Sample 3 | Sample 4 |
|---|---|---|---|---|
| **CCL17 (pg/ml)** | 7.3 | 2 | 31.2 | <LLOD |
| **Dilution Factor used** | 1/10 | 1/10 | 1/10 | 1/100 |
| **Elastase-1 (mg/ml)** | 119.03 | 8.75 | 1.94 | 6.73 |
| **Dilution Factor used** | 1/100,000 | 1/65,000 | 1/8,500 | 1/40,000 |
| **IL-8 (mg/ml)** | 379.99 | 31.73 | 17.12 | 29.97 |
| **Dilution Factor used** | 1/120,000 | 1/90,000 | 1/30,000 | 1/85,000 |
| **IP-10 (mg/ml)** | 11.4 | 80.96 | 6.75 | 177.3 |
| **Dilution Factor used** | 1/10,000 | 1/8,000 | 1/7,500 | 1/90,000 |
| **SAA (ng/ml)** | 3.91 | 6467.6 | 13.94 | <LLOD |
| **Dilution Factor used** | 1/10 | 1/9,500 | 1/10 | 1/8,000 |
| **MxA (pg/ml)** | 148.75 | 4922.84 | 109.45 | 891.42 |
| **Dilution Factor used** | 1/20 | 1/100 | 1/20 | 1/100 |

**Supplementary Table 3: Descriptive statistics of protein recovery in samples spiked with low and high concentrations of protein.**

| | Low Spiking Concentration | | | High Spiking Concentration | | |
|---|---|---|---|---|---|---|
| **Cytokine** | Mean (%) | SEM (%) | CV (%) | Mean (%) | SEM (%) | CV (%) |
| **CCL17** | 72.1 | 8.9 | 24.8 | 82.6 | 17.9 | 43.3 |
| **Elastase-1** | 103.8 | 3.9 | 7.6 | 100.3 | 2.2 | 4.4 |
| **IL-8** | 100.6 | 3.9 | 7.7 | 92.3 | 0.38 | 0.82 |
| **IP-10** | 127.0 | 13.9 | 21.9 | 99.5 | 5.9 | 11.8 |
| **SAA** | 93.4 | 4.7 | 10.0 | 79.4 | 2.3 | 5.7 |
| **MxA** | 105 | 9.1 | 17.4 | 83.9 | 4.6 | 10.9 |

**Supplementary Table 4: CCL17 protein concentration detected in neat and spiked nasal samples.**

| CCL17 (pg/ml) | Neat | After Low Spike | After High Spike |
|---|---|---|---|
| **Sample 1** | 7.30 | 5 | 576.78 |
| **Sample 2** | 2.00 | 3 | 323 |
| **Sample 3** | 31.20 | 7 | 586 |
| **Sample 4** | <LLOD | 5 | 965 |

**Supplementary Table 5: Elastase-1 protein concentration detected in neat and spiked nasal samples.**

| Elastase-1 (ng/ml) | Neat | After Low Spike | After High Spike |
|---|---|---|---|
| **Sample 1** | 119234 | 264699 | 602296 |
| **Sample 2** | 8756 | 89051 | 332819 |
| **Sample 3** | 1944 | 12490 | 44614 |
| **Sample 4** | 6795 | 59246 | 221846 |

**Supplementary Table 6: IL-8 protein concentration detected in neat and spiked nasal samples.**

| IL-8 (ng/ml) | Neat | After Low Spike | After High Spike |
|---|---|---|---|
| **Sample 1** | 376196 | 1025237 | 6966340 |
| **Sample 2** | 41113 | 621901 | 5022982 |
| **Sample 3** | 13704 | 197068 | 1674327 |
| **Sample 4** | 32618 | 540953 | 4797469 |

**Supplementary Table 7: IP-10 protein concentration detected in neat and spiked nasal samples.**

| IP-10 (mg/ml) | Neat | After Low Spike | After High Spike |
|---|---|---|---|
| **Sample 1** | 11.40 | 144 | 142.21 |
| **Sample 2** | 80.96 | 381.1 | 379.91 |
| **Sample 3** | 6.75 | 219.72 | 237.56 |
| **Sample 4** | 177.30 | 657.47 | 776.27 |

**Supplementary Table 8: SAA protein concentration detected in neat and spiked nasal samples.**

| SAA (pg/ml) | Neat | After Low Spike | After High Spike |
|---|---|---|---|
| **Sample 1** | 3909.2 | 7653.91 | 56112.67 |
| **Sample 2** | 6467600.0 | 20186.97 | 21674.4 |
| **Sample 3** | 13941.1 | 9011.91 | 55789.99 |
| **Sample 4** | <LLOD | 2402.06 | 1769.45 |

**Supplementary Table 9: MxA protein concentration detected in neat and spiked nasal samples.**

| MxA (ng/ml) | Neat | After Low Spike | After High Spike |
|---|---|---|---|
| **Sample 1** | 0.1 | 11 | 44 |
| **Sample 2** | 4.9 | 52 | 228 |
| **Sample 3** | 0.1 | 12 | 37 |
| **Sample 4** | 0.9 | 41 | 181 |
